# Trans-ethnic eQTL meta-analysis of human brain reveals regulatory architecture and candidate causal variants for brain-related traits

**DOI:** 10.1101/2021.01.25.21250099

**Authors:** Biao Zeng, Jaroslav Bendl, Roman Kosoy, John F. Fullard, Gabriel E. Hoffman, Panos Roussos

**Affiliations:** Pamela Sklar Division of Psychiatric Genomics, New York, New York, USA; Department of Genetics and Genomic Sciences, New York, New York, USA; Icahn Institute for Data Science and Genomic Technology, New York, New York, USA; Department of Psychiatry, Icahn School of Medicine at Mount Sinai, New York, New York, USA; Mental Illness Research, Education and Clinical Centers, James J. Peters VA Medical Center, Bronx, New York, USA

## Abstract

While large-scale genome-wide association studies (GWAS) have identified hundreds of loci associated with neuropsychiatric and neurodegenerative traits, identifying the variants, genes and molecular mechanisms underlying these traits remains challenging. Integrating GWAS results with expression quantitative trait loci (eQTLs) and identifying shared genetic architecture has been widely adopted to nominate genes and candidate causal variants. However, this integrative approach is often limited by the sample size, the statistical power of the eQTL dataset, and the strong linkage disequilibrium between variants. Here we developed the multivariate multiple QTL (mmQTL) approach and applied it to perform a large-scale trans-ethnic eQTL meta-analysis to increase power and fine-mapping resolution. Importantly, this method also increases power to identify conditional eQTL’s that are enriched for cell type specific regulatory effects. Analysis of 3,188 RNA-seq samples from 2,029 donors, including 444 non-European individuals, yields an effective sample size of 2,974, which is substantially larger than previous brain eQTL efforts. Joint statistical fine-mapping of eQTL and GWAS identified 301 variant-trait pairs for 23 brain-related traits driven by 189 unique candidate causal variants for 179 unique genes. This integrative analysis identifies novel disease genes and elucidates potential regulatory mechanisms for genes underlying schizophrenia, bipolar disorder and Alzheimer’s disease.

## Introduction

Genome-wide association studies (GWAS) have associated hundreds of loci with neuropsychiatric and neurodegenerative traits (Jansen et al., 2019; Nalls et al., 2019; Schizophrenia Working Group of the Psychiatric Genomics Consortium, 2014; Visscher et al., 2017; Wray et al., 2018). Yet elucidating the molecular mechanisms underlying these traits remains challenging since most risk variants are non-coding and highly correlated due to linkage disequilibrium (Schaid et al., 2018; Visscher et al., 2017). Integration of risk loci with expression quantitative trait loci (eQTL) has been widely adopted to identify genes and candidate causal variants (Gallagher and Chen-Plotkin, 2018; GTEx Consortium, 2020; Hormozdiari et al., 2016). Recent work by the Genotype-Tissue Expression (GTEx) consortium across 838 individuals and 49 tissues, detected eQTLs for 95% of protein-coding and >60% of long non-coding RNA genes (GTEx Consortium, 2020). While the power to detect primary (i.e. the most significant association) eQTLs is very high, advances in identifying tissue- and cell-type-specific effects, conditionally independent effects, and candidate causal variants in trait-relevant tissues and cell types promises to further inform the molecular etiology of disease (Dobbyn et al., 2018; GTEx Consortium, 2020; Hormozdiari et al., 2016, 2018; Kim-Hellmuth et al., 2020).

Large-scale efforts have been undertaken to catalogue human brain eQTLs (Fromer et al., 2016; GTEx Consortium, 2020; Jaffe et al., 2018; Ng et al., 2017; Wang et al., 2018a). All these efforts focus on homogenate brain tissue, which is composed of multiple cell types (Cao et al., 2020; Darmanis et al., 2015; Habib et al., 2017; Lake et al., 2018), and, therefore, cell type-specific eQTLs are not fully captured (Fairfax et al., 2014; Raj et al., 2014; van der Wijst et al., 2018). This is an important limitation given that disease variants act through cell-type-specific biological effects (Farh et al., 2015; Finucane et al., 2018; Raj et al., 2014). Initial efforts have performed cell type-specific eQTL analysis in human brain by experimentally purifying specific cell types (Jaffe et al., 2020; de Paiva Lopes et al., 2020; Young et al., 2019), but the sample size of such studies are necessarily limited by the increased experimental costs, and data quality can be affected by the additional experimental steps. An alternative strategy to capture cell type-specific effects is to statistically define conditional- or context-dependent eQTL (Dobbyn et al., 2018; Kim-Hellmuth et al., 2020). While existing studies have sufficient power to detect primary eQTLs, identifying conditionally independent eQTLs that capture more subtle cell type-specific effects requires large sample sizes (Jansen et al., 2017; Zhernakova et al., 2017).

Following eQTL detection, statistical fine-mapping can identify candidate causal variants likely to drive variation in expression (Benner et al., 2016; Hormozdiari et al., 2014, 2016; Schaid et al., 2018). Going one step further, joint statistical fine-mapping integrating GWAS and gene expression traits can define the candidate causal variants that increase disease risk through alterations of gene expression (Hormozdiari et al., 2016). Interpreting and validating such variants can pinpoint genes such as *FURIN* (Schrode et al., 2019), *BIN1* (Nott et al., 2019) and *C4* (Sekar et al., 2016) along with molecular mechanisms that can be further studied in experimental systems. Yet the resolution of statistical fine-mapping for eQTL and GWAS is incomplete due to limited sample sizes and lack of trans-ancestry analysis (Schaid et al., 2018). Sample size of more than 2,000 donors is needed to detect eQTLs and perform GWAS colocalization for identification of causal variants explaining 1% of heritability (Hormozdiari et al., 2016). The largest current human brain eQTL mega-analysis by PsychENCODE included 1,387 unique donors from multiple cohorts (Wang et al., 2018a). Moreover, most eQTL analyses have been limited to European populations, despite the fact that much shorter linkage disequilibrium in individuals of African or African-American ancestry can substantially increase the resolution of statistical fine-mapping (Asimit et al., 2016; Morris, 2011; Schaid et al., 2018; Zaitlen et al., 2010).

Given the limited availability of human brain samples, it is critical to maximize power and fine-mapping resolution by combining existing datasets. Yet differences in study designs have, thus far, hindered such efforts. Trans-ethnic studies have long been challenging in genetics, but linear mixed models can control the false positive rate in the presence of complex population structure (Sul et al., 2018; Yang et al., 2014; Zhou and Stephens, 2012). Moreover, expression measurements from multiple brain regions in GTEx are not statistically independent, so combining these data entails explicit modelling of these correlated measurements from the same set of individuals (Han et al., 2016).

In order to realize the potential of trans-ethnic eQTL fine-mapping and integration with brain-related GWAS results, we developed the multivariate multiple QTL (mmQTL) pipeline and applied it to a combined analysis of brain tissues from PsychENCODE, Religious Orders Study and Memory and Aging Project (ROSMAP) and GTEx. Our pipeline performs eQTL detection with a linear mixed model, identifies conditionally independent eQTL and combines results across datasets with a random effects meta-analysis that models the correlation between multiple brain regions from a shared set of individuals. Joint fine-mapping then identifies candidate causal variants shared between gene expression and GWAS traits. This integrative analysis identifies novel disease genes and elucidates potential regulatory mechanisms for genes underlying schizophrenia (SZ), bipolar disorder (BD) and Alzheimer’s disease (AD).

## Results

### Analysis overview

We performed a trans-ethnic eQTL meta-analysis on RNA-seq gene expression data from non-overlapping samples from the dorsolateral prefrontal cortex (DLPFC) from PsychENCODE (Wang et al., 2018a) and ROSMAP (Bennett et al., 2018), and 13 brain regions from GTEx (GTEx Consortium et al., 2017) (**Figure 1A**). We accounted for diverse ancestry (**Figure 1B**), repeated measures from a shared set of donors in GTEx and effect size heterogeneity using a linear mixed model for analysis of each dataset, followed by combining these 15 eQTL analyses using a random effects meta-analysis (**Figure 1C**). This statistical framework is implemented in our mmQTL software (**see Methods**). Statistical fine-mapping of the eQTL meta-analysis was integrated with GWAS fine-mapping from CAUSALdb (Wang et al., 2020) to identify candidate causal variants shared between gene expression and neuropsychiatric traits.

**Figure 1:**
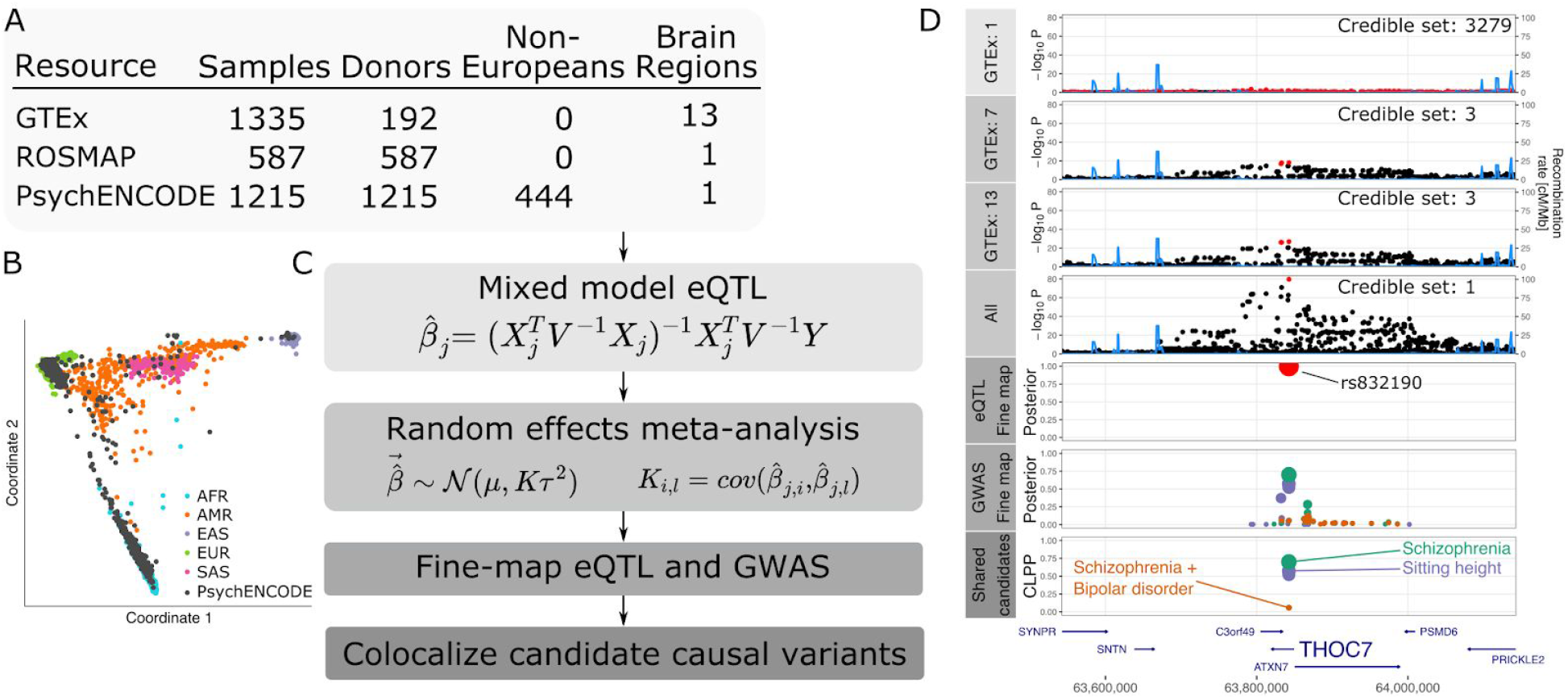
Workflow for trans-ethnic eQTL meta-analysis. **A)** RNA-seq datasets with details about ancestry and repeated measures. **B)** Multidimensional scaling illustrating diverse ancestry of donors from PsychENCODE resource. **C)** mmQTL workflow is composed of eQTL analysis within each brain region for each resource using a linear mixed model to account for population stratification. Each analysis is then combined using a random effects meta-analysis that accounts for repeated measures from GTEx sample and effect size heterogeneity across brain regions and resources. Statistical fine-mapping is performed on GWAS and combined eQTL results separately. Finally, fine-mapping posterior probabilities from the eQTL analysis and each GWAS are combined to produce <u>c</u>o<u>l</u>ocalization <u>p</u>osterior <u>p</u>robabilities (CLPP). **D)** Analysis of data for *THOC7* from 1, 7 and 13 GTEx brain tissues, and addition of PsychENCODE and ROSMAP, reduces the size of the 95% credible sets indicated by red points. Statistical fine-mapping for this gene and integration with GWAS nominates a single candidate causal variant, rs832190, affecting SZ, a combined risk for SZ and BD, and sitting height in this region.

For example, results for *THOC7* illustrate that increasing the number of GTEx tissue from 1 to 7 to 13 enhances power and decreases the size of the 95% credible sets, while integration with PsychENCODE and ROSMAP nominates a single candidate causal variant (**Figure 1D**). Integrating GWAS and eQTL results produces colocalization posterior probabilities (CLPP) > 0.05 for SZ, BD and sitting height, and identifies rs832190 and *THOC7* as the candidate causal variant and gene, respectively, for this locus.

### Biologically motivated simulations

Simulations motivated by the scenarios considered here (i.e. diverse ancestry and repeated measures design of the human brain datasets) were used to evaluate mmQTL performance in terms of: 1) controlling the false positive rate, 2) leveraging eQTL effects shared across multiple tissues and 3) reducing the size of the credible set from statistical fine-mapping (**Figure 2**). For the eQTL analysis we considered a linear regression model including 5 genotype PC’s and a linear mixed model that counts for the genetic similarity between all pairs of samples (Sul et al., 2018; Yang et al., 2014; Zhou and Stephens, 2012). The summary statistics for each SNP-gene pair were aggregated across tissues using a fixed- or random-effects meta-analysis, or simply the minimum p-value with a Sidak correction to account for the number of tissues. The first two explicitly account for the repeated measures design by modeling the correlation between summary statistics under the null, while the Sidak-corrected minimum p-values assume independence.

**Figure 2:**
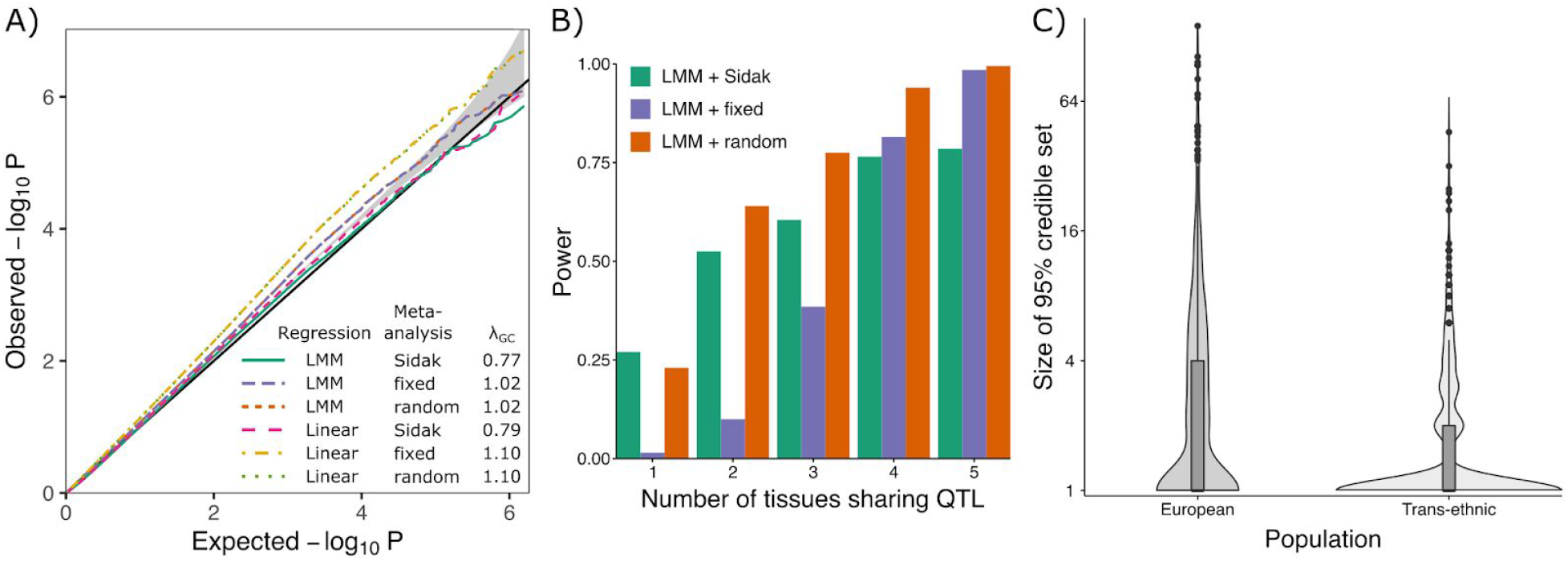
Biologically motivated simulations demonstrate performance of mmQTL workflow: high correlation scenario. **A)** QQ plot of results from null simulation shows that the linear mixed model (LMM) with fixed or random effect meta-analysis accurately controls the false positive rate, while linear regression with 5 genotype principal components did not. The Sidak method was very conservative in both cases. λ_GC_ indicates the genomic control inflation factor. **B)** Power from LMM followed by 3 types of meta-analysis versus the number of tissues sharing an eQTL. **C)** Size of the 95% credible sets from statistical fine-mapping for a dataset of European samples versus a trans-ethnic dataset of the same size.

We simulated genotypes for 500 individuals in each of three distinct populations: European, African, and Asian. A single causal eQTL explaining 1-2% of expression variation in up to 5 tissues for these 1,500 individuals was simulated for 800 randomly chosen genes where the number of tissues with a shared effect varied from 1 to 5. Correlation between the same gene expression trait measured in two tissues was simulated to be low (r=0.12) or high (r=0.45) (**see Methods**).

In a null simulation with all genetic effects set to zero in both the low and high correlation scenarios, the linear mixed model accurately controlled the false positive rate when summary statistics from multiple tissues were aggregated using the Sidak method as well as fixed or random effects meta-analysis (**Figure 2A, Supplementary Figure 1**). As expected, the linear model did not adequately account for the complex population structure and showed an inflated false positive rate. Therefore, it was not included in subsequent simulations.

Power analyses were performed on the same set of samples of diverse ancestry where the number of tissues with a shared eQTL effect varied between 1 and 5 (**Figure 2B**). Using a p-value cutoff of 10^−6^, the random effects meta-analysis following a linear mixed model eQTL analysis had the highest power under most levels of eQTL sharing across tissues because it models heterogeneity in effect sizes across tissues. The fixed-effect meta-analysis was less powerful because it assumes a shared effect size across tissues. The Sidak corrected minimum p-value only performed best when the eQTL was tissue-specific (i.e. no cross-tissue sharing) since it assumes statistical independence of the results from each tissue.

The mmQTL workflow with linear mixed model followed by a random-effects meta-analysis demonstrated accurate control of the false positive rate while retaining high power under biologically motivated simulations. With the goal of identifying candidate causal variants shared with brain-related traits, we evaluated the benefit of using a dataset of diverse ancestry. A dataset of 1,500 European individuals was simulated in addition to the trans-ethnic cohort above. One causal variant with effect size was used to simulate gene expression traits. Statistical fine-mapping of eQTL results from the trans-ethnic cohort produced 95% credible sets containing a mean of 2.0 SNP’s compared to a mean of 4.8 for the European only cohort (**Figure 2C**). In the trans-ethnic cohort, 73.0% of genes have a single candidate causal variant compared to 51.6% in the European cohort. Moreover, random-effects meta-analysis reduces the credible set by 10.0% compared to fixed effects meta-analysis.

### Evaluating mmQTL workflow on real data

Here we evaluate the empirical performance of our mmQTL workflow on real data by analysing an increasing number of brain regions (k=1,4,7,13) from GTEx (**Figure 3**). As expected, mmQTL is able to borrow information across multiple brain regions using a random-effects meta-analysis so that increasing k substantially increases the empirical effective sample size (N_eff_) (**Figure 3A**). With k=13, there are 1,335 RNA-seq samples from 192 individuals producing empirical N_eff_ = 524. Moreover, increasing k decreases the median size of the 95% credible sets from statistical fine-mapping (**Figure 3B**).

**Figure 3:**
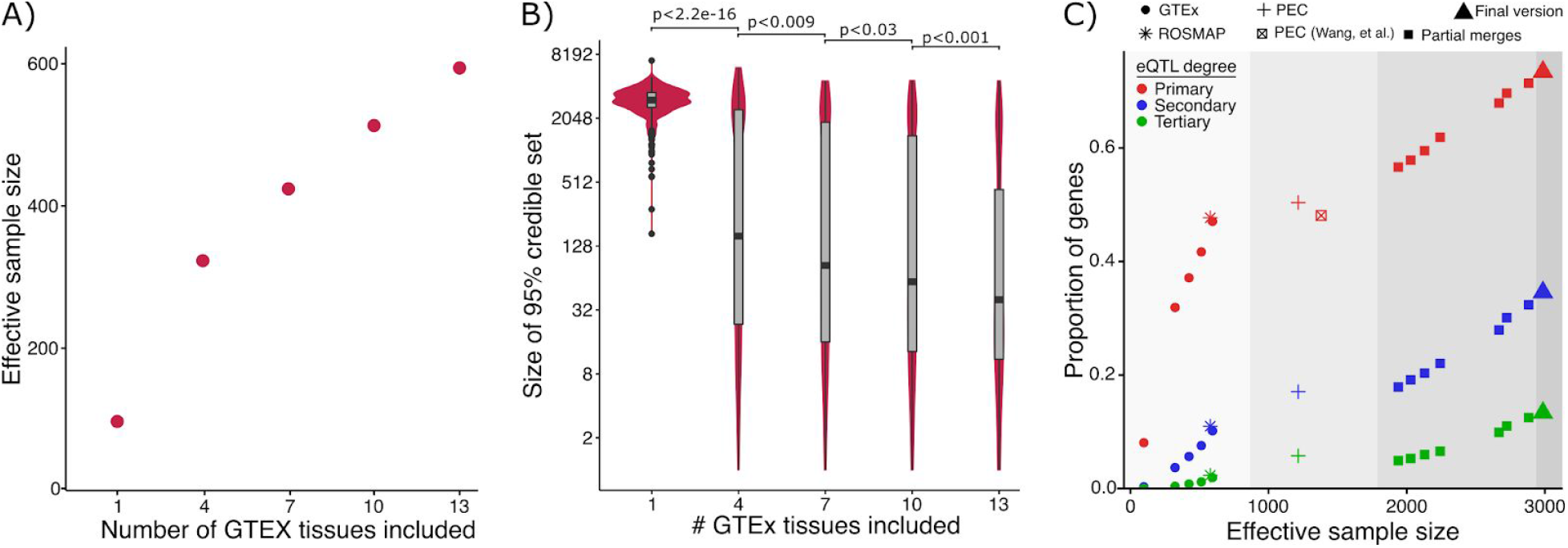
Evaluation of mmQTL workflow on real data. **A)** Increasing the number of brain regions from GTEx increases the effective sample size. **B)** Increasing the number of brain regions from GTEx decreases the median 95% credible set size. P-values are shown from one-sided Kolmogorov–Smirnov test between adjacent categories. **C)** Including additional datasets increases the proportion of genes with a detectable primary or conditional eQTL. Colors indicate degree of eQTL. Panel is divided into regions showing 1) GTEx and ROSMAP results; 2) PsychENCODE (PEC) data analyzed here, and published PEC summary statistics (Wang et al., 2018a); 3) adding an increasing number of GTEx brain tissues to the PEC+ROSMAP results; 4) final version merging PEC+ROSMAP+GTEx.

The value of adding each successive study to the meta-analysis was evaluated for primary eQTLs as well as secondary and tertiary conditional eQTLs using a conservative p-value cutoff of 10^−6^ (**Figure 3C, see Methods**). The PsychENCODE study included the largest cohort and yielded 50.4% of genes having genome-wide significant primary eQTLs. Adding data from GTEx and ROSMAP produced a combined eQTL analysis comprising 3,188 RNA-seq samples from 2,029 donors to give N_eff_ = 2,974. Powered by this substantial increase in N_eff_, eQTLs were detected for 73% of genes analysed in the final meta-analysis.

### Properties of brain eQTL meta-analysis

Our brain eQTL meta-analysis identifies 10,456 genes with a genome-wide significant eQTL, including 4,808 with at least one conditional eQTL using a conservative p-value threshold of 10^−6^ (**Figure 4A**). These eQTL results are highly reproducible with estimated replicated rate π_1_=75.6% when evaluated in an independent dataset of bulk brain tissue (Wang et al., 2018b) using Storey’s π_1_ statistic (Storey and Tibshirani, 2003). The increased power from our meta-analysis enables detection of cell-type-specific eQTL not detectable in smaller studies of bulk brain tissue. eQTLs detected in the granule cell layer of the dentate gyrus enriched for excitatory neurons (Jaffe et al., 2020), are replicated in our analysis at π_1_=83.8% compared to π_1_=65.2% in the PsychENCODE analysis (one-sided z-test p < 1.15e^-8^), and eQTLs detected in purified microglia (Kosoy, et al, in preparation) are replicated in our analysis at π_1_=89.0% compared to π_1_ =55.0%, from the PsychENCODE analysis (one-sided z-test p < 6.24^-6^) (**Figure 4B**). Overlaying variants in 95% credible sets with with ATAC-seq regions identified by fluorescence activated nuclei sorting for 4 cell populations (GABAergic neurons, glutamatergic neurons, oligodendrocytes, and a mixture of microglia and astrocytes) (Hauberg et al., 2020) identifies significant enrichment within open chromatin regions for each cell population (**Figure 4C**). Conditional eQTLs have different properties than primary eQTLs. While primary eQTLs are a median of 24 kb from the transcription start site, conditional eQTL are more distal with median distances of 39 kb for secondary, 53 kb for tertiary and 75 kb for quaternary eQTL (p < 0.001 for all comparisons of adjacent categories using one sided Kolmogorov–Smirnov test) (**Supplementary Figure 2A**). This is consistent with primary eQTLs often affecting promoters and conditional eQTL more often affecting enhancers. In addition, genes with more independent eQTLs have higher cell type specificity in human (Darmanis et al., 2015) (Spearman rho = 0.0408, p = 1.85×10^−6^) and mouse (Zeisel et al., 2015) (Spearman rho: 0.04435, p = 2.18×10^−7^) brain (**Supplementary Figure 2B)**. Finally, genes with more conditional eQTLs tend to be under lower evolutionary constraint, as measured by the probability of loss intolerance (pLI) calculated from large-scale exome sequencing (Lek et al., 2016). While 35% of genes with no detectable eQTLs are highly constrained (pLI > 0.9), only 10% of genes with 4 eQTLs exceed this cutoff (**Supplementary Figure 2C)**.

**Figure 4:**
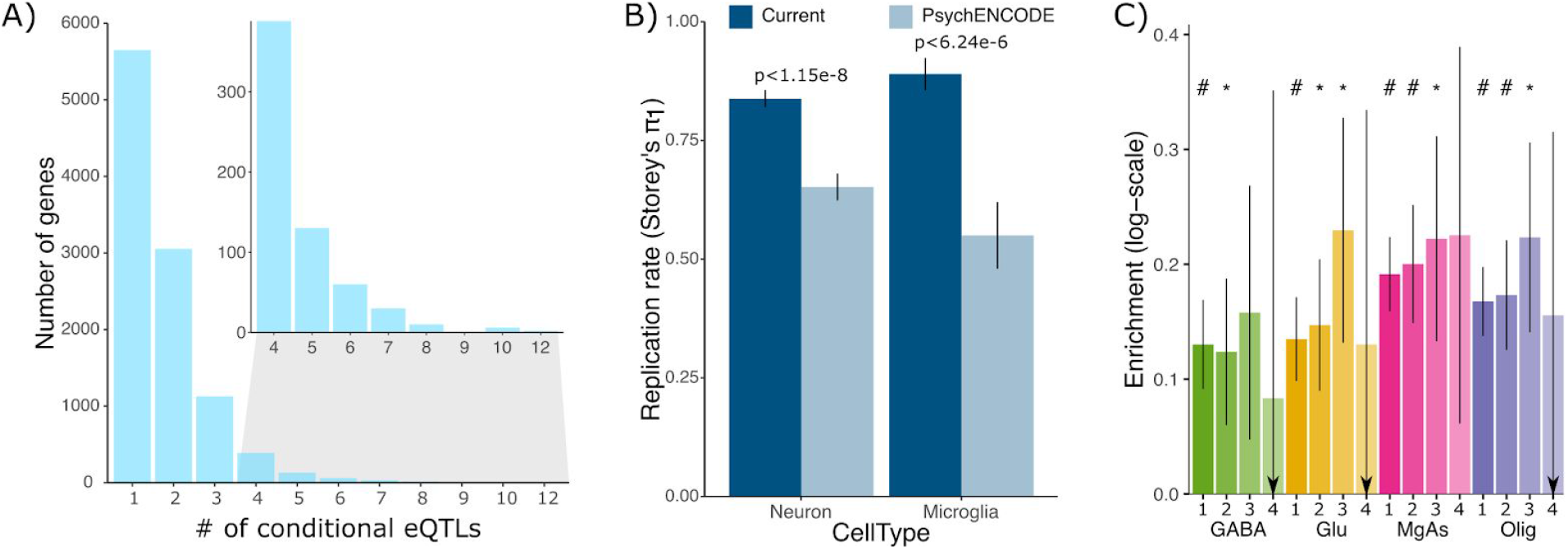
Properties of brain eQTL meta-analysis. **A)** Number of genes having a significant primary or conditional eQTL for degree up to 12. Inset shows number of genes for eQTL degree 4 to 12. **B)** Replication rate measured by Storey’s π_1_ in the current study and PsychENCODE for eQTLs discovered in the granule cell layer of the dentate gyrus enriched for excitatory neurons (Jaffe et al., 2020), and purified microglia (Kosoy, in preparation). Error bar indicates standard error from 100 bootstrap samplings. P-value indicates one-sided z-test. **C)** Enrichment of variants in the 95% causal sets for each gene in open chromatin regions assayed in each of 4 cell populations. Results are shown for eQTL degree 1 to 4. Error bars indicate standard deviation, ‘#’ indicates Bonferroni adjusted p-value < 0.05 and ‘*’ indicates nominal p-value < 0.05.

### Variants in credible sets are enriched for risk for brain-related traits

Integration of variants in the 95% credible set for primary and conditional eQTLs with large-scale GWAS summary statistics using stratified linkage disequilibrium scores regression (Finucane et al., 2015) finds significant enrichments across 22 complex traits after accounting for baseline annotations (**Figure 5**). Variants in the 95% credible set for primary eQTLs were enriched for 21 traits, including 8 neuropsychiatric and behavioral traits, and 4 neurodegenerative diseases. Meanwhile, the enrichment for conditional eQTLs was limited to AD, BD and alcohol use. These enrichments indicate that our meta-analysis and statistical fine-mapping captures risk variants for brain-related phenotypes.

**Figure 5:**
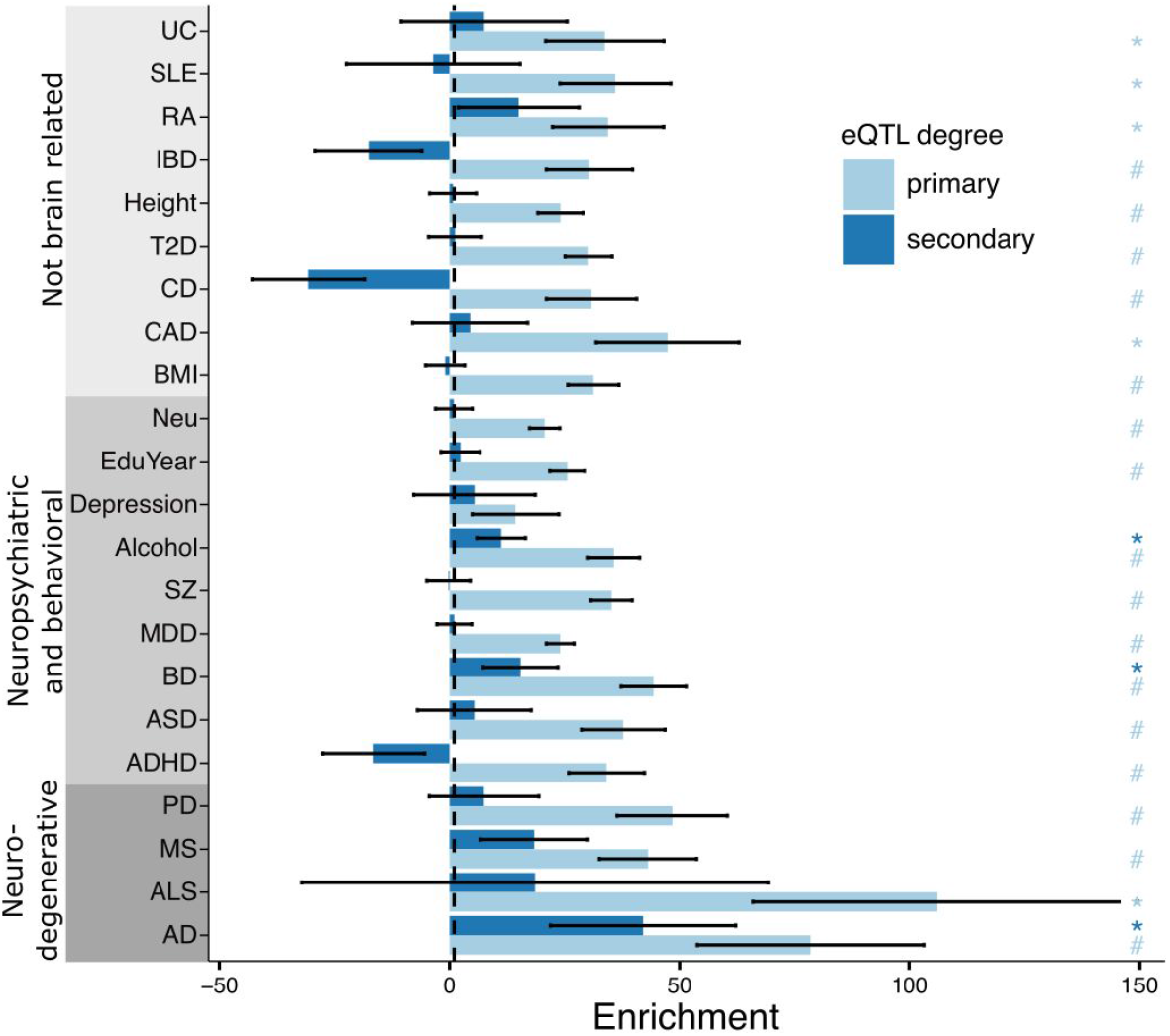
Heritability enrichment of variants in the 95% causal set for 22 complex traits. Linkage disequilibrium score regression (LDSC) enrichments are shown for variants in the 95% causal set for primary and secondary eQTLs. Error bars indicate standard errors. ‘#’ indicates p-value passes 5% Bonferroni cutoff for 44 tests and ‘*’ indicates p-value < 0.05. See Supplementary Table 1 for trait abbreviations and references.

### Fine mapping identifies candidate causal variants conveying risk for brain-related traits

Integrating our eQTL fine-mapping results with candidate causal variants from large-scale GWAS (Wang et al., 2020) using a joint fine-mapping approach (Hormozdiari et al., 2016) identifies 6,978 variant-trait pairs (CLPP > 0.01) including 2,048 unique candidate causal variants and 1626 unique genes among 683 complex traits (**Supplementary Figure 3**). These results include 301 variant-trait pairs for 23 brain-related traits for 189 and 179 unique candidate causal variants and genes, respectively (**Figure 6A**). Analysis of SZ and BD, two neuropsychiatric diseases with high genetic co-heritability (Bipolar Disorder and Schizophrenia Working Group of the Psychiatric Genomics Consortium, 2018; Cross-Disorder Group of the Psychiatric Genomics Consortium., 2019; Pardiñas et al., 2018) identified candidate causal variants for 20 genes predicted to confer risk for one or both diseases (**Figure 6B**). The top genes with CLPP > 0.5 for either of these diseases include *ZNF823, THOC7* and *FURIN*. While these genes have been implicated in SZ or BP previously, and in fact the candidate causal variant for *FURIN*, rs4702, has been validated experimentally (Schrode et al., 2019), candidate causal variants for the other two genes have not been previously identified. Moreover, integrating results from analysis of SZ, BP and SZ+BP versus controls indicates the specificity of these candidates causal variants. *ZNF823* is predicted to confer risk to SZ, but not BD. *THOC7* has a substantially larger CLPP score for SZ compared to the joint SZ+BP GWAS. Conversely, *FURIN* has a higher CLPP for the joint SZ+BP GWAS than for SZ alone. Notably, the candidate causal variants driving the colocalization with SZ and BD for *CACNA1C* and *VARS2* are in fact secondary eQTLs, emphasizing the importance of including conditional eQTL analysis. The candidate causal variant for *CACNA1C*, rs2007044, which is within an intronic enhancer, has been previously shown to affect transcription due to reduced promoter interaction (Roussos et al., 2014).

**Figure 6:**
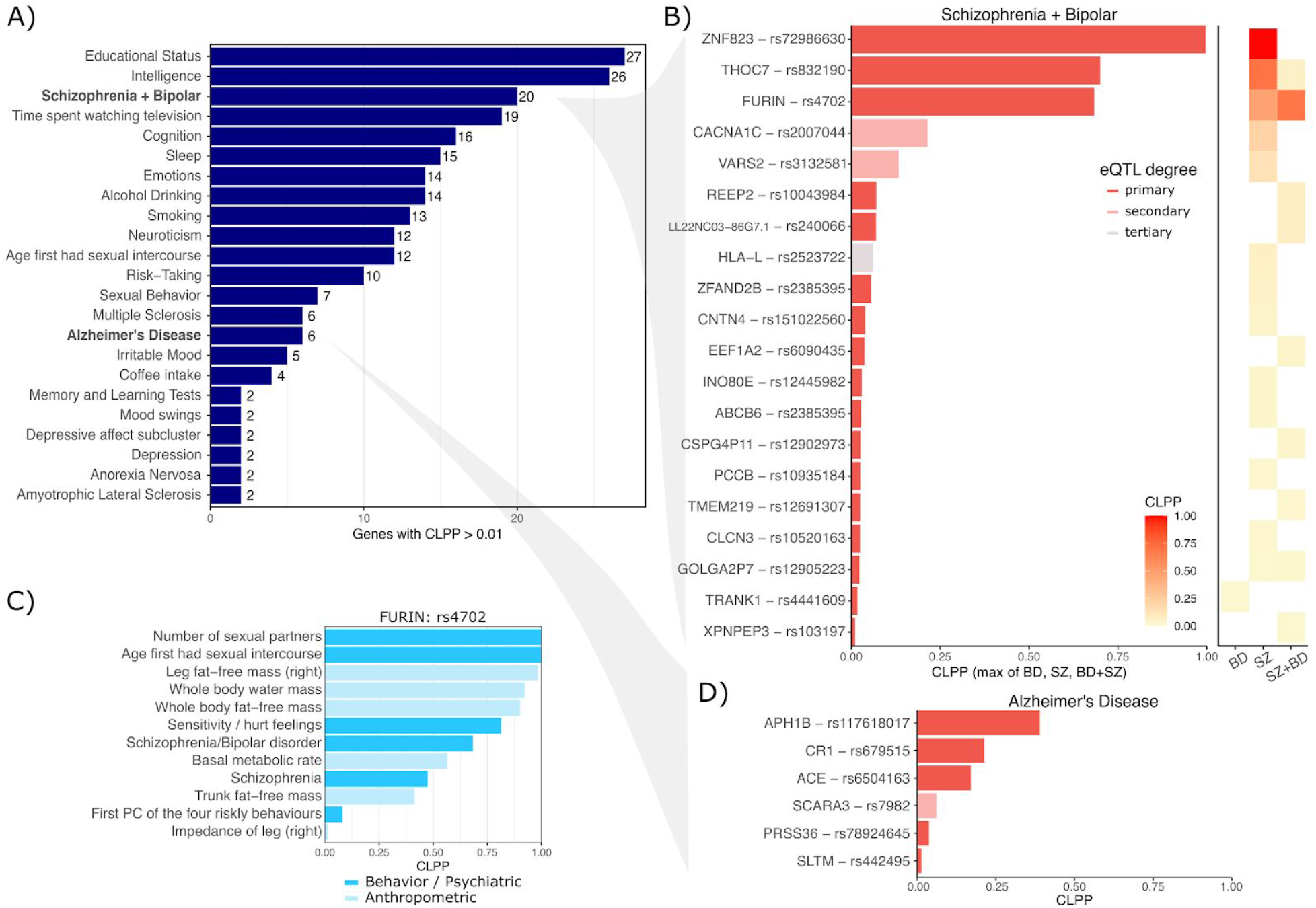
Summary of joint fine-mapping colocationation with brain-related traits. **A)** Number of genes with colocalization posterior probability (CLPP) > 0.01 for **B)** Genes with CLPP > 0.01 for Schizophrenia (SZ) and Bipolar Disorder (BP) and a joint GWAS of SZ+BP versus controls. For each gene, the max CLPP across SZ, BP and SZ+BP is shown. Right Panel shows CLPP for BP, SCZ and SZ+BP compared to controls. **C)** A validated casual variant, rs4702, that affects expression of *FURIN* is predicted to affect risk for multiple complex behavioral, psychiatric and anthropometric traits. **D)** Genes with CLPP > 0.01 for AD.

In addition, analysis of candidate causal variables across many phenotypes enables insight into pleiotropy. *FURIN* and rs4702 are also implicated in the number of sexual partners, age at first sexual intercourse, risk taking behavior, and emotional sensitivity / hurt feelings, and multiple anthropometric traits (**Figure 6C, Supplementary Figure 4**). Sharing of a candidate causal variant and gene between SZ+BP and these risk-taking behavior traits is particularly interesting given that impulsiveness is a clinical feature of both SZ and BD (Najt et al., 2007; Ouzir, 2013), and is associated with more severe psychiatric symptoms and decreased level of functioning (Cerimele and Katon, 2013).

Analysis of AD identified candidate causal variants for 6 genes. While these genes have been highlighted previously (Jansen et al., 2019), our analysis highlights variants and their mechanistic link to disease (**Figure 6D)**.

### Candidate causal variants elucidate potential molecular mechanisms

rs117618017 is the top causal variant for AD and drives the expression of *APH1B*, a subunit of the gamma-secretase complex, which includes multiple AD risk genes as components (**Figure 7A**). This missense coding variant was identified in a GWAS meta-analysis for AD (Jansen et al., 2019), but an attempt to experimentally validate a functional effect from this single amino acid change yielded only negative results (Zhang et al., 2020). Yet our analysis indicates an alternative molecular mechanism, whereby, instead of acting by changing protein sequence, the minor allele of rs117618017 increases AD risk by directly increasing gene expression of *APH1B*.

**Figure 7:**
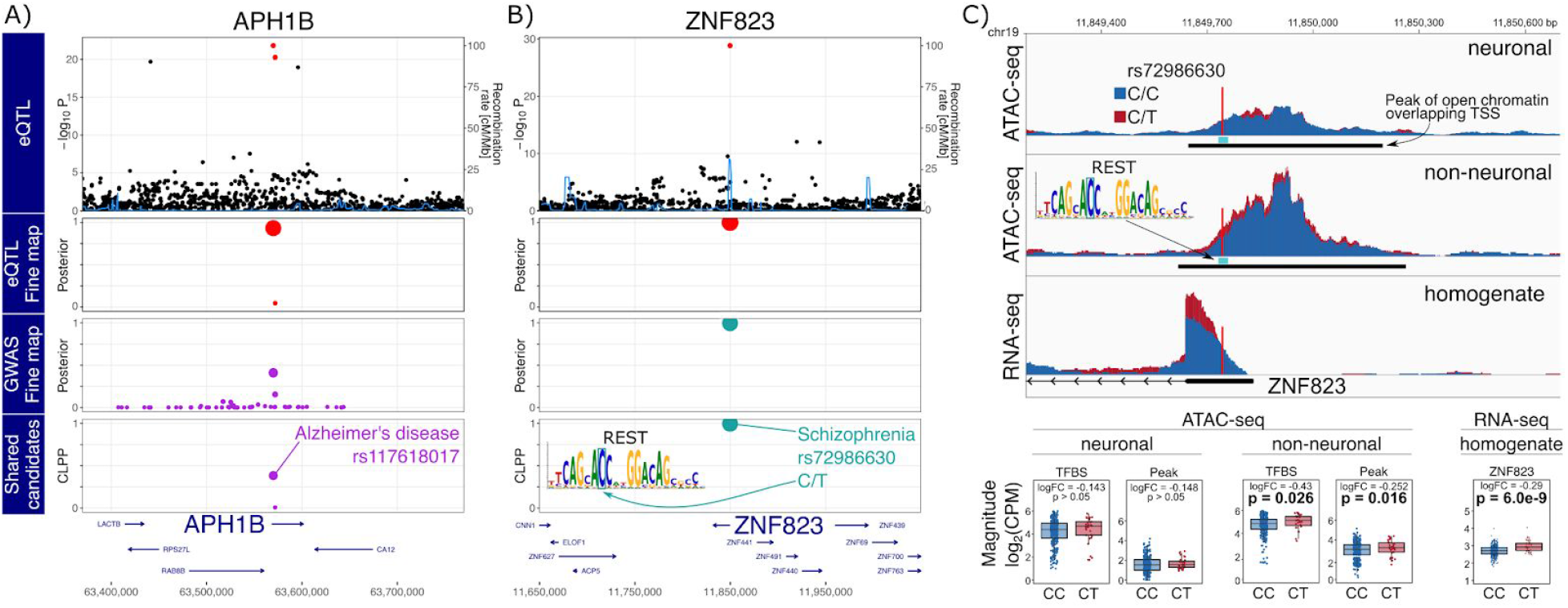
GWAS-eQTL colocationation by joint fine-mapping. **A**,**B)** Starting from the top, the plot shows -log_10_ p-values from eQTL analysis, poster probabilities from statistical fine-mapping of eQTL results, poster probabilities from statistical fine-mapping of GWAS results, and colocalization posterior probabilities (CLPP) for combining eQTL and GWAS fine-mapping. **A)** Expression of *APH1B* and AD risk share rs117618017 as a candidate causal variant. **B)** Expression of *ZNF823* and SZ risk share rs72986630 as a candidate causal variant. This variant is predicted to disrupt a REST binding site motif. **C)** Individuals heterozygous for rs72986630 have increased chromatin accessibility at the peak and REST binding site in non-neuronal cells. Genome-plot shows chromatin accessibility for neuronal (top) and non-neuronal (middle) nuclei, and gene expression from brain homogenate bottom. The lower panel shows boxplots comparing chromatin accessibility and gene expression between individuals with two reference alleles (i.e. CC) compared to CT heterozygotes.

The top hit for SZ is rs72986630, which is predicted to drive expression of *ZNF823*, a zinc finger protein with little additional annotation (**Figure 7B**). This C/T SNP is located in the 5’ UTR of the gene and the minor allele, T (MAF ∼6%), is protective against SZ. This variant is predicted to disrupt a binding site for the RE1 silencing transcription factor (REST), also known as neuron-restrictive silencing factor. REST is upregulated during neurogenesis and in adult non-neuronal cells, and acts by silencing neuron specific genes (Hwang and Zukin, 2018; Schoenherr and Anderson, 1995). Analysis of chromatin accessibility in this region using a large-scale ATAC-seq dataset from purified neuronal and non-neuronal nuclei from the anterior cingulate cortex (ACC) of post mortem brains of 368 donors elucidated the molecular mechanism (Bendl, et al. in preparation) (**Figure 7C**). In non-neuronal cells, but not in neuronal cells, individuals heterozygous at this site have higher chromatin accessibility at both the 644 bp ATAC-seq peak (p = 0.016) and the 21 bp motif (p = 0.026), and this corresponds to decreased binding of REST at this site. Since REST is a transcriptional silencer, decreased binding of REST should lead to increased expression of *ZNF823*. Querying RNA-seq data from brain homogenate from these samples confirms that heterozygous individuals have increased expression of *ZNF823* (p=6.01×10^−9^).

## Discussion

Integration of eQTL and GWAS is a powerful method to understand the molecular mechanism influencing complex traits. While transcript-wide association studies aim to identify genes underlying a complex trait, correlated expression and co-regulation can be challenging to overcome (Mancuso et al., 2019; Wainberg et al., 2019). Joint fine-mapping focuses instead on identifying variants that drive both gene expression and a downstream trait (Hormozdiari et al., 2016). Despite recent successes, fine-mapping is often limited by statistical power and linkage disequilibrium (Hormozdiari et al., 2016; Schaid et al., 2018). Our mmQTL workflow addresses both of these issues by performing a trans-ethnic eQTL meta-analysis of 3,188 RNA-seq samples from 2,029 donors, with an effective sample size of 2,974, to produce the largest resource to date characterizing the genetics of gene expression in the human brain. This analysis has substantially boosted the catalog of genes with detected conditional eQTLs, while increasing the resolution of statistical fine-mapping.

Despite being performed on bulk RNA-seq data, our analysis is able to replicate eQTLs discovered in purified microglia (Kosoy, in preparation) and neurons (Jaffe et al., 2020), and the replication rate is substantially higher than for PsychENCODE (Wang et al., 2018a). Moreover, we identify candidate causal variants enriched in cell type specific open chromatin regions. While much recent work has pursued generating eQTLs from purified cell populations (Jaffe et al., 2020; de Paiva Lopes et al., 2020; Young et al., 2019), and eQTL discovery from single cell/nucleus RNA-seq is becoming tractible (Mandric et al., 2020; van der Wijst et al., 2020), our eQTL meta-analysis from bulk tissue illustrates thats large sample size and sophisticated statistical modelling has substantial power to replicate eQTLs from smaller studies of purified cell types.

While the number of genes with detectable eQTLs approaches saturation, there is substantial value in increasing sample size. Here, we use individuals of diverse ancestry paired with a linear mixed model in our mmQTL workflow to increase the resolution of statistical fine-mapping. Moreover, we perform conditional eQTL analysis to identify genes with up to 12 independent eQTLs. These conditional eQTLs tend to have smaller effect sizes, be farther from transcription start sites, and affect genes that are more cell type specific. The number of genes with secondary and tertiary eQTL does not appear close to saturation, underscoring the regulatory variation that remains to be identified.

Integrating statistical fine-mapping for eQTLs and GWAS across hundreds of complex traits enabled insight into candidate causal variants, mechanisms of disease genetics and pleiotropy. Focusing on regulatory mechanisms for genes underlying brain-related traits, we identified 20 genes and candidate causal variants predicted to drive risk for SZ and BD, plus another 6 for AD. While other methods focus on discovering disease genes, here we focus on discovering gene-variant pairs underlying disease risk in order to elucidate the molecular mechanisms that convey risk.

Here we highlighted two examples. The SNP rs117618017 is a candidate causal variant causing a single amino acid change in *APH1B*. While experimental results of the impact of this amino acid change were negative (Zhang et al., 2020), our analysis instead supports a mechanism where this variant increases disease risk by increasing expression of APH1B. Our analysis predicts that rs72986630 drives expression of *ZNF823* and is protective against SZ. By integrating chromatin accessibility data from post mortem brains, we traced the predicted chain of causality and found that the minor allele disrupts binding of REST in non-neuronal cells, which then increases expression of *ZNF823*. The lack of an effect in neuronal nuclei is consistent with the higher expression of REST in non-neuronal cells during adulthood, silencing neuron-specific genes (Hwang and Zukin, 2018; Schoenherr and Anderson, 1995).

While we focused on regulatory mechanisms for genes underlying SZ, BD and AD, all results are available from the Brain eQTL meta-analysis (BREMA) resource (icahn.mssm.edu/brema).

Further integration of multi-omics data with trans-ethnic fine-mapping and large-scale GWAS promises to yield further insight into the molecular mechanisms underlying disease risk. Future studies are poised to perform multiple genomic assays, namely RNA-seq and ATAC-seq, on multiple tissues or brain regions, and target multiple cell types either by sorting or single cell/nucleus methods (Mandric et al., 2020; van der Wijst et al., 2020). Moreover, these studies will increasingly include individuals of diverse ancestry (Wojcik et al., 2019). Our mmQTL method will enable the field to take advantage of these repeated measures datasets while modeling effect size heterogeneity and controlling the false positive rate. Efforts to trace the chain of causality from variants and molecular mechanisms to pleiotropy across complex phenotypes are poised to yield insight into novel therapeutic targets.

## Data Availability

All results are available at http://icahn.mssm.edu/brema

http://icahn.mssm.edu/brema

## Resources

Brain eQTL meta-analysis resource: http://icahn.mssm.edu/bremam

mQTL: https://github.com/jxzb1988/mmQTL

## Contributions

B.Z., G.E.H. and P.R. conceived and designed the study; B.Z. designed and implemented the statistical method; B.Z and G.E.H. performed the analysis; J.F.F. generated the cell type specific expression and chromatin accessibility data; J.B. and R.K. preprocessed and analyzed cell type specific expression and chromatin accessibility data; J.F.F. and P.R. supervised the data generation; G.E.H. and P.R. supervised the data analysis; G.E.H., B.Z. and P.R. wrote the manuscript with the help of all authors.

## Funding

The project was supported by the National Institute of Mental Health, NIH grants R01-MH109677, U01-MH116442, R01-MH125246 and R01-MH109897, the National Institute on Aging, NIH grants R01-AG050986, R01-AG067025 and R01-AG065582, and the Veterans Affairs Merit BX004189 to P.R.. G.E.H. was supported in part by NARSAD Young Investigator Grant 26313 from the Brain & Behavior Research Foundation. J.B. was supported in part by NARSAD Young Investigator Grant 27209 from the Brain & Behavior Research Foundation. Research reported in this paper was supported by the Office of Research Infrastructure of the National Institutes of Health under award numbers S10OD018522 and S10OD026880. The content is solely the responsibility of the authors and does not necessarily represent the official views of the National Institutes of Health.

## Methods

### Obtaining and processing of RNA-seq datasets

Imputed genotypes from GTEx v7 were downloaded from dbGAP (Accession phs000424.v7.p2). For ROSMAP, the imputed genotypes were downloaded from the Synapse website (id: syn3157329). The imputed genotype for each cohort in the PsychENCODE study was downloaded from Synapse website (id: syn21052530), and was then filtered to retain variants with imputation quality ≥0.3. Filtered genotypes from each cohort were merged and variants with MAF ≥ 1% and Hardy-Weinberg Equilibrium p-value ≥ 1e-6 were retained.

The original PsychENCODE analysis performed eQTL detection using 1,387 individuals (Wang et al., 2018a). In the current work we exclude a small fraction of these individuals. First, the full PsychENCODE dataset contains GTEx samples which we excluded in order to avoid redundancy with our separate GTEx analysis. Second, the original analysis used ∼5 million imputed SNPs. Since accurate statistical fine-mapping depends on including the true causal variant in the analysis, we included additional well-imputed SNPs at the cost of excluding a small set of samples. Excluding samples with < 8 million well-imputed (info score >0.3) variants yielded 1,215 individuals used in this study.

The normalized gene expression of GTEx v7 was downloaded from GTEx Portal (GTEx_Analysis_2016-01-15_v7_RNASeQCv1.1.8_gene_tpm.gct.gz, https://gtexportal.org/), and we regressed out covariates from the companion file GTEx_Analysis_v7_eQTL_covariates.tar.gz with linear regression. Normalized data from PsychENCODE (`DER-01_PEC_Gene_expression_matrix_normalized.txt`) was downloaded from http://resource.psychencode.org/, and as the downloaded gene expression is already normalized regressing out the effect of covariates, no further normalization was taken. Data from ROSMAP (syn3388564, `ROSMAP_RNAseq_FPKM_gene.tsv`) was downloaded from https://adknowledgeportal.synapse.org. The provided FPKM abundance values were quantile normalized, log2 transformed, standardized to normal distribution, and 20 principal components of the gene expression matrix were regressed out.

### Linear mixed model eQTL analysis

Given expression abundance of a gene measured in tissues from same set of individuals, the gene expression in tissue can be modeled as:

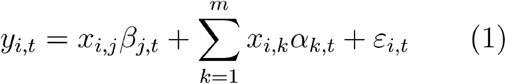

where, *y*_*i,t*_ is the measured gene expression value for individual *i* in tissue which has been normalized so that it has mean 0 and variance 1,*x_i,j_* is the genotype dosage for individual *i* at variant *j* normalized so that it has mean 0 and variance 1,*β*_*j,t*_ is effect size for variant *j* and *t* tissue. The next term models the polygenic background across *m* variants where x_*i,k*_ is the genotype dosage value for individual *i* at variant *k* and *α*_*k,t*_is the effect size for variant *k*and tissue *t* with distribution 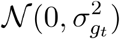, where 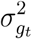 is the tissue-specific parameter for genetic background. Finally, *ε*_*i,t*_ is the normally distributed error variance for individual *i* and tissue *t* with distribution 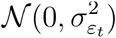, where 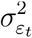 is the tissue-specific parameter for random noise.

This linear mixed model can be transformed for practical estimation the effect size *β*_*j,t*_ Equation (1) can be rewritten as

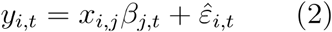

where 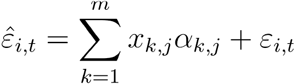 and has a distribution 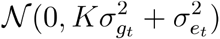, where *K* is a genetic relatedness matrix estimated based on genome-wide - genotypes.

Considering that the phenotype was collected among *l* individuals, we can write formula (2) into a vector format:

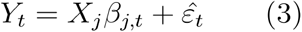

where *Y*_*t*_,*X*_*i*_ and 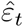 are *l*-dimensional vectors, and contain normalized phenotype, normalized genotype of variant *j*, and noise, respectively.

From Equation (3), *β*_*j,t*_ can be estimated as

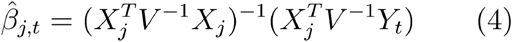

Where 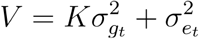 and produces an unbiased estimator since

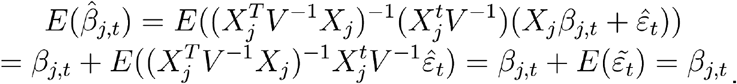

### Modeling covariance across tissues

While standard meta-analysis assumes that effect size estimates are statistically independent, analysis of multiple tissues from the same set of subjects produces covariance between the coefficient estimates. Here we explicitly model this covariance in order to control the false positive rate.

Denote the estimate for variant *j* across all tissues as the vector 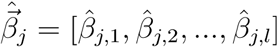 Since individuals overlap across the multiple tissues, the entries of 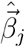 will be correlated. Estimating coefficients for tissues 1 and 2 using Equation 4 gives

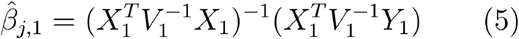

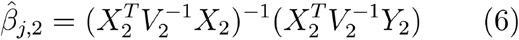

where an index is added to to distinguish the two tissues which may have partial sample overlapping. These estimates are not statistically independent since

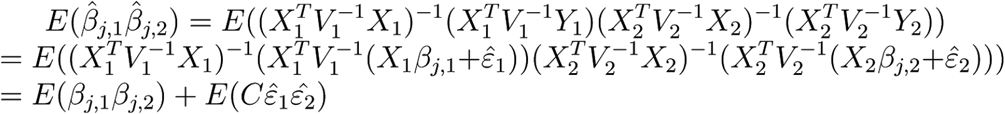

Where 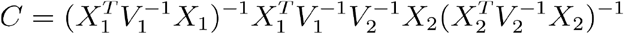 is only involved with transformed genotypes projected by a covariance matrix. Noting that 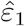 and 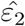 are the summed contribution from polygenic background and noise, if there are sample overlapping and the phenotypes share causal variants in two tissues, then

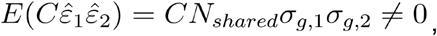

Where N_*shared*_ is the number of shared individuals, and σ_*g*,1_ and σ_*g*,2_ are the genetic component for polygenic background in tissue 1 and tissue 2. Finally we note that

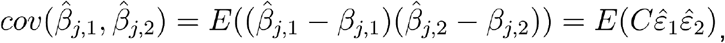

explicitly indicating that there is nonzero covariance between estimators. Our mmQTL method estimates the covariance matrix among *n* tissues based on the non-significant z-score in tissues, and set it to be *Ĉ*. This matrix is defined so that the covariance between tissues *i* and *j* is estimated by the covariance between z-scores from non-significant variants (p>0.05) according to:

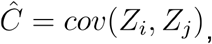

Where *Z*_*i*_ and *Z*_*j*_ are vectors containing statistical Z-scores.

### Fixed-and random effects meta-analysis

The results from multiple analyses are aggregated using either a fixed or random effects meta-analysis. The true effects sized are assumed to be drawn from a normal distribution 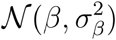 centered at the true effect size *β* with variance 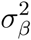. For a fixed effect model, the true effect size is fixed at a constant value which is equivalent to setting 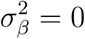 and for the random effects model 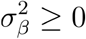. From this hierarchical framework, we obtain estimators for variant *j* among tissues, denoted as a vector 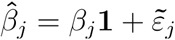 which has a distribution 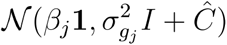. We applied the Brent-method implemented in C++ Boost library (https://www.boost.org) to estimate *β*_*j*_ and 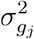. To test the difference with null hypothesis, we applied the random-effect model2 (Han and Eskin, 2011; Han et al., 2016) to obtain a p-value.

### Detection of conditional eQTLs

We applied a stepwise selection strategy explore cis-region and identify conditionally independent eQTL associations. An iterative strategy is applied to find conditional independent eQTL: previously detected eQTL signals are regressed out and another round of eQTL detection was initiated. If one or more variants with p-value less than 10^−6^, the variant with the smallest p-value is added to the list of conditionally independent effects. The process is repeated until no addition variant has a p-value < 10^−6^. If a high-order eQTL is in high LD with low-order eQTL (r^2^>=0.3), the high-order eQTL will be excluded in order to avoid attenuating the estimated effect size of low-order eQTL.

Importantly, we demonstrate statistically that the order in which conditional eQTLs are detected is biologically meaningful: large-effect eQTL shared among tissues are likely to be detected first, while and small-effect eQTL or tissue-specific effect will be detected as higher-order eQTL.

Consider two true causal variants *i* and where *j* the estimated effect has the distribution around the true value according to 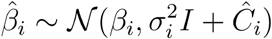, where *Ĉ*_*i*_ is defined above. The non-centrality parameter (NCP) reflecting the statistical power for this variant is

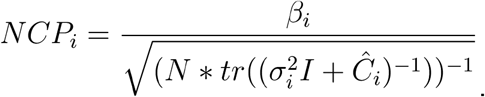

The ratio between the NPC of variant *i* and the second variant *j* is

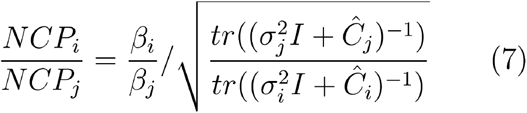

From empirical observation that the effect size (with the genotype and response normalized) of primary eQTL is much larger than that of non-primary eQTL, *Ĉ*_*i*_ ≈ *Ĉ*_*j*_ and both are positive definite, and can be decomposed *U*Σ*U*, in which *U* consists of the eigenvectors, and Σis a diagonal matrix with elements being eigenvalues, denoted as *diag*(λ_1_, λ_2_, λ_3_, …, λ_*N*_) 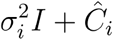 can be decomposed into 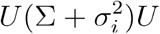, and 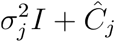to be 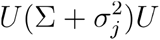.

Therefore, Equation (7) can be rewritten as

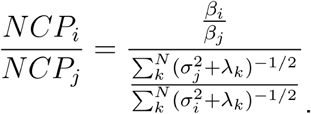

Based on this, it is apparent that the difference in statistical power for variants *i* and *j* is mainly determined by effect size, and its variance. For a variant with larger effect size, and smaller variance, it has a higher statistical power, which is consistent with the empirical that primary eQTL has a much larger normalized effect size (*β*_i_ > *β*_*j*_), and smaller variance because of the sharing among tissues 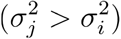, so 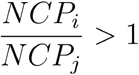. It follows that on average, mmQTL will pick the independent eQTL signal in a biologically meaningful manner, so that eQTL with a larger influence on expression abundance among conditions tends to be selected first.

### Multiple testing

Multiple testing correction is performed at the level of the locus as well as genome-wide. Empirically, we performed the locus-level control applying Bonferroni correction, which is a most conservative strategy, and Benjamini-Hochberg method (Benjamini and Hochberg, 1995) on genome-wide correction, and we found that a p-value 10^−6^ is enough for two-level multiple test correction. While studies often use more liberal multiple testing cutoffs because of the limited statistical power, the statistical fine-mapping that is the focus of this analysis can perform poorly on genes that only pass a liberal cutoff (Hormozdiari et al., 2016, 2018).

### Computing empirical effective sample size

In linear regression model for QTL analysis, given that both phenotype and genotype were normalized, the estimator for the allelic effect size is 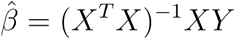, and its variance is 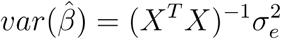. Letting 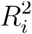 be the variance explained by the explored variant and *N*_*i*_ be the (effective) sample size for study *i*, the variance of the effect size estimate is 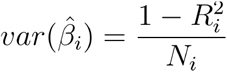. Consider two studies, where the (effective) sample size of the first study is easy to estimate just by using the number of samples, and the second has some confounding factors such as repeat measurements or population structure. Assuming that the effect size of a given causal variant is constant in the two studies, the ratio of the variances is determined only by *N*_1_ and *N*_2_ :

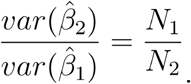

Therefore the effective sample size, *N*_2_, can be computed from known values by 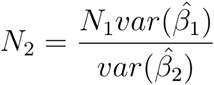.

We used individual brain tissue in GTEx dataset as study 1 to define *N*_1_, and eQTL results from fixed-effect meta-analysis as study 2. The genome-wide variance ratio was set to be the median ratio of variances based on all variants with abs(z-score)>=10 in the fixed-effect meta analysis. When evaluating the effective size of a meta-analysis, the effective sample size was computed by treating each brain tissue in GTEx as the baseline study and then taking the mean estimate effective sample size over 13 brain regions.

### Replication of eQTLs from purified cell types

In order to assess the replication of eQTLs discovered in independent datasets, we considered the lead SNP for each gene with a genome-wide significant eQTL in the granule cell layer of the dentate gyrus enriched for excitatory neurons (Jaffe et al., 2020) and purified microglia (Kosoy, et al., in preparation). For the set of lead SNPs from each dataset, the p-values were extracted from the current eQTL analysis as well as the PsychENCODE analysis (Wang et al., 2018a) and Storey’s π_1_ was evaluated using qvalue (Storey and Tibshirani, 2003). The PsychENCODE p-values were obtained from http://resource.psychencode.org/Datasets/Derived/QTLs/Full_hg19_cis-eQTL.txt.gz. Uncertainty in π_1_ estimates were evaluated using 100 bootstraps where SNPs were sampled with replacement and π_1_ was recomputed each time. A p-value comparing the replication rate for the current and PsychENCODE analysis was computed using a one-sided z-test using the estimated π_1_ values and their bootstrap variances.

### Simulation pipeline to evaluate mmQTL performance

Genotype and gene expressions data simulated to compare to empirical performance of eQTL analysis using a linear model with 5 genotype principal components compared to a linear mixed model. Results from eQTL analysis of 5 simulated tissues were then aggregated using either Sidak correction, or a fixed- or random-effects meta-analysis.

Biologically realistic genotype data reflecting real human populations was simulated with a sampling-based simulation package, hapgen2 (Su et al., 2011), and haplotype information for European, African, and Asian populations from the 1000 Genomes Project (https://mathgen.stats.ox.ac.uk/impute/data_download_1000G_2010_interim.html). We simulated 500 individuals for each population, and merged these individuals into a single trans-ancestry dataset with sample size 1,500. We also simulated 1,500 individuals solely based on European haplotype information.

Based on these genotypes, we adapted the phenotypesimulator pipeline (Meyer and Birney, 2018) to perform 800 simulations for each scenario, simulating one gene expression trait for each simulation. For each gene a single eQTL was simulated to affect expression abundance explaining 1% phenotypic variance, and the contribution due to polygenic background was set to be 30%. We applied phenotypesimulator’s simulating strategy to account for shared environmental factors, measurement noise, and polygenic background to create correlated phenotypes. In the simulation, we simulated phenotypes in 5 tissues, and set the number of tissues that the causal genetic variant affects to be 1, 2, 3, 4, 5. To demonstrate the robustness of mmQTL to control for population structure and batch effect, we set two different levels of phenotype correlation, a low level, r=0.12, and the high level r=0.45. For power analysis, any simulated causal variants located in high LD (r^2^ >= 0.8) with a variant passing the multiple testing cutoff was considered to be detected.

We also performed a null simulation with no true causal variants where all effect sizes were set to zero. Results from 50 simulations were aggregated and we used genomic inflation factor (Devlin and Roeder, 1999) and QQ plots to assess the false positive rate.

Comparison of fine-mapping resolution between a European and trans-ethnic population was performed using simulated pure 1,500 European European individuals and trans-ethnic 1,500 individuals with 500 individuals in each of European, African and Asian population. Gene expression phenotype was simulated in a single tissue, and a causal variant was randomly chosen to explain 2% phenotypic variance. For 1,500 European individuals, we applied standard linear regression model to detect eQTL and then fine-mapping was conducted to obtain a 95% credible set candidate for causal variants, while for 1,500 trans-ethnic individuals, we used mixed linear model to detect eQTL and then fine-mapping was taken to find a 95% credible set. The size of the 95% credible set was used to compare the fine-mapping resolution, a smaller number indicating a higher fine-mapping resolution.

### Integration with ATAC-seq data

Variants in the 95% credible set were overlaid with open chromatin regions from four distinct populations of cells (glutamatergic neurons, GABAergic neurons, oligodendrocytes, and a mixture of microglia/astrocytes) identified by ATAC-seq (Hauberg et al., 2020). In order to reduce the influence of the low fine-mapping resolution of conditional eQTL, if the size of the 95% credible set for a single gene contained >10 variants, only the 10 variants with highest PIP were included. Enrichment of variants within open chromatin regions was evaluated using a Fisher’s exact test implemented in QTLTools (Delaneau et al., 2017).

### Evaluating GWAS enrichments for variants in credible sets

We applied a strategy developed in Hormozdiari et al. (Hormozdiari et al., 2018): for each eQTL, we performed fine-mapping and compute the causal posterior probability (CPP) of each cis-SNP and only variants in the fine-mapped 95% credible set are left for following analysis. For each SNP in cis-regions, we assign an annotation value based on the maximum value of CPP across all molecular phenotypes; SNPs that do not belong to any 95% Credible Set are assigned an annotation value of 0, which is referred as MaxCPP in (Hormozdiari et al., 2018). Stratified linkage disequilibrium (LD) score regression (S-LDSC) (Finucane et al., 2015) was then used to partition trait heritability using the constructed functional annotations, and the estimated enrichment was used to measure the importance of each eQTL category on human complex traits or diseases. To rule out the potential influences of the correlation among eQTL categories, we aggregate the baselineLD model, which includes a set of 75 functional annotations, and functional annotations for eQTL categories and run S-LDSR simultaneously.

GWAS summary statistics were obtained for 22 human complex traits or diseases, which contain both brain traits and non-brain traits (**Supplementary Table 1)**. The summary results were firstly converted into the required format for LDSR by the provided `“munge_sumstats.py”` command in the LDSC package (https://github.com/bulik/ldsc).

### eQTL detection in cell type specific datasets

Microglia from fresh human brain specimens (101 samples, including 27 non-Europeans) were prepared using the Adult Brain Dissociation Kit (Miltenyi Biotech). Tissue homogenates were incubated in antibody (CD45: BD Pharmingen, Clone HI30 and CD11b: BD Pharmingen, Clone ICRF44) at 1:500 for 1 hour in the dark at 4°C with end-over-end rotation. Prior to FACS, DAPI (Thermoscientific) was added at 1:1000 to facilitate identification of dead cells. Viable (DAPI negative) CD45^**+**^/CD11b^**+**^ cells were isolated by FACS using a FACSAria flow cytometer (BD Biosciences) (Kosoy et al., in preparation). RNA was extracted from FACS sorted cells (Arcturus PicoPure RNA Isolation Kit, Life Technologies) and sequencing libraries generated using the SMARTer Stranded RNA-seq kit (Clontech), according to manufacturer’s instructions. Variants with MAF > 5%, and Hardy-Weinberg equilibrium test p-value > 10^−6^ were retained and analyzed using a linear mixed model implemented in mmQTL. Gene expression was normalized using log2 CPM and eQTL analysis was performed on residauals after regression out 15 principal components of the gene expression. For each gene, a Benjamini-Hochberg (BH) FDR correction was applied across all variants tested in the cis regulatory region to obtain the minimum q-value. Then, the minimum q-values across all genes are adjusted again by the BH FDR method to compute the genome-wide FDR. Limited by the small sample size, we chose a less conservative FDR cutoff of 10%.

### Statistical fine-mapping

For each detected eQTL, we conducted a fine-mapping analysis applying the CAVIAR method (Hormozdiari et al., 2014) implemented in mmQTL to find a 95% credible set for causal variants. Briefly, meta-analysis p-value based on a random-effect model in each round of conditional eQTL detection was firstly converted to z-score, which was then used as input for fine-mapping. CAVIAR will calculate the posterior inclusion probability (PIP) of each variant to causal, and a set of variants prioritized by PIP score were outputted with summed PIP equal to 0.95.

### Detecting colocalization between eQTL and GWAS signals

Joint statistical fine-mapping of eQTL’s and GWAS signals (Hormozdiari et al., 2016) was performed by multiplying the estimated posterior inclusion probability (PIP) for a given variant from the eQTL analysis by the PIP for this variant from GWAS of traits compiled in CausalDB (Wang et al., 2020) to obtain a <u>c</u>o<u>l</u>ocalization <u>p</u>osterior <u>p</u>robability (CLPP). A gene is considered to share a candidate causal variant with a GWAS trait if at least one variant has a CLPP > 0.01 (Hormozdiari et al., 2016).

### Trait classification

CausalDB (Wang et al., 2020) provided the MeSH Category for each GWAS trait. However, brain-related traits fall in multiple MeSH categories and there is no single criterion to identify such traits. We performed manual inspection of traits in CausalDB that could be considered neuropsychiatric, neurodegenerative or behavioral and termed them ‘brain related’.

### Validation of rs72986630 effect in chromatin accessibility and gene expression data

To further investigate one prioritized functional variant rs72986630 that reside in REST TF binding site overlapping TSS of *ZNF823*, we queried our unpublished ATAC-seq data set (Bendl et al., in preparation) of neuronal and non-neuronal samples from ACC brain region generated on postmortem human brains from CommonMind cohort (Hoffman et al., 2019). This dataset consists of samples from 370 donors (114 SZ cases, 64 BD cases, 64 controls) with rs72986630 MAF of 6.0%. Since only two donors carry the ALT/ALT (i.e. T/T) genotype, we excluded them for further analysis.

To generate ATAC-seq data set, neuronal and non-neuronal cell populations were isolated from postmortem tissue by fluorescence-activated nuclear sorting using anti-NeuN antibody. ATAC-seq libraries were created using an established protocol (Buenrostro et al., 2015). Raw sequencing reads were mapped to human genome hg38 using STAR aligner (Dobin et al., 2013). The samples of the same cell type (neuron / non-neuron) and genotype at rs72986630 (CC / CT) were subsampled and merged, creating 4 BAM files with a uniform depth of 1 billion pair-end reads. Subsampling ratios were calculated per each sample individually within those four respective groups (genotype by cell type) to ensure that each of them contributed the same number of reads, regardless of their per-sample read counts. Using these BAM files, bigWig files were created and peaks were called by the MACS (v2.1) with the same parameters as described in (Hauberg et al., 2020), but using an FDR threshold of 0.01. After removing peaks overlapping the blacklisted genomic regions and peaks not being sufficiently accessible (CPM>1 in at least 10% of samples was required), 498,183 peaks remained. The final read count matrix of 664 samples by 498,183 peaks was normalized using the trimmed mean of M-values (TMM) method. The following covariates were selected by Bayesian Information Criterion (BIC) method to be added to the base covariates, i.e. genotype by cell type: mean GC content, fraction of reads with GC content 0-19%, 20-39%, 40-59%, fraction of reads in peaks, fraction of unmapped reads, AT dropout, and mean insert size. Since our dataset contains up to two samples per individual, we ran differential analysis to get differentially accessible peaks between CC and CT carriers using dream (v1.17.9) (Hoffman and Roussos, 2020) that accounts for correlation structure in repeated measures. As an alternative approach, instead of quantifying changes between all open chromatin regions, we performed differential analysis between CC and CT carriers on TF binding sites of REST motif. We used footprinting to narrow down our focus only to 31,534 REST TF binding sites that are bound in at least one set of samples (out of 4 sets, i.e. genotype by cell type) as predicted by TOBIAS (Bentsen et al., 2020).

The analysis of differential gene expression between REF/REF (C/C) and REF/ALT (C/T) genotype at rs72986630 followed the same approach as the analysis of chromatin accessibility. We used a subset of 338 homogenate RNA-seq samples of ACC brain region from CommonMind Consortium (Hoffman et al., 2019) that originate from the same donors as ATAC-seq samples. We performed differential analysis only for sufficiently expressed protein-coding genes (CPM>1 in at least 30% of samples was required), i.e. we start the analysis with a count matrix of 338 samples by 14,893 genes that were normalized by trimmed mean of M-values (TMM) method. The following technical covariates were selected by BIC method: institution, expression profiling efficiency, intronic rate, intragenic rate, fraction of reads with GC content 20-39%, 40-59%, and AT dropout.

## Supplementary Figures

**Supplementary Figure 1:**
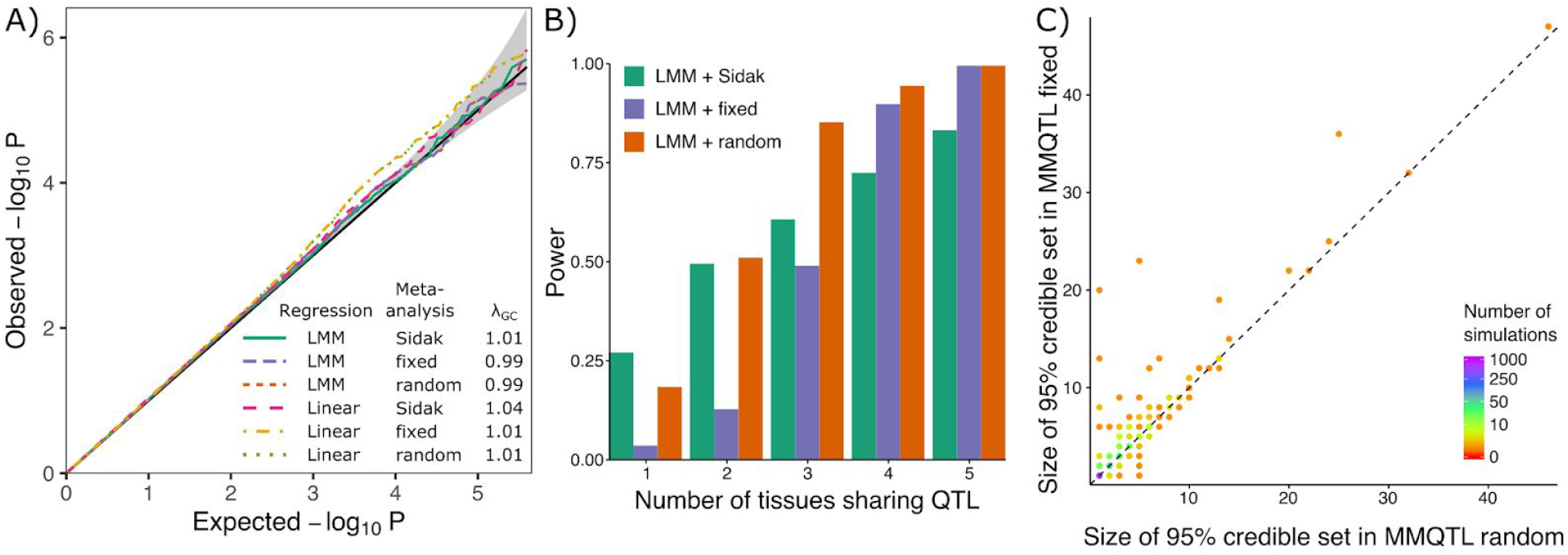
Biologically motivated simulations demonstrate performance of mmQTL workflow: low correlation scenario. **A)** QQ plot of results from null simulation shows that the linear mixed model (LMM) with fixed or random effect meta-analysis accurately controls the false positive rate for, while linear regression with 5 genotype principal components did not. The Sidak method was very conservative in both cases. λ_GC_ indicates the genomic control inflation factor. **B)** Power from LMM followed by 3 types of meta-analysis versus the number of tissues sharing an eQTL. **C)** Size of the 95% credible sets from fixed-(y-axis) and random-(x-axis) effects meta-analysis from simulations in Figure 2C.

**Supplementary Figure 2:**
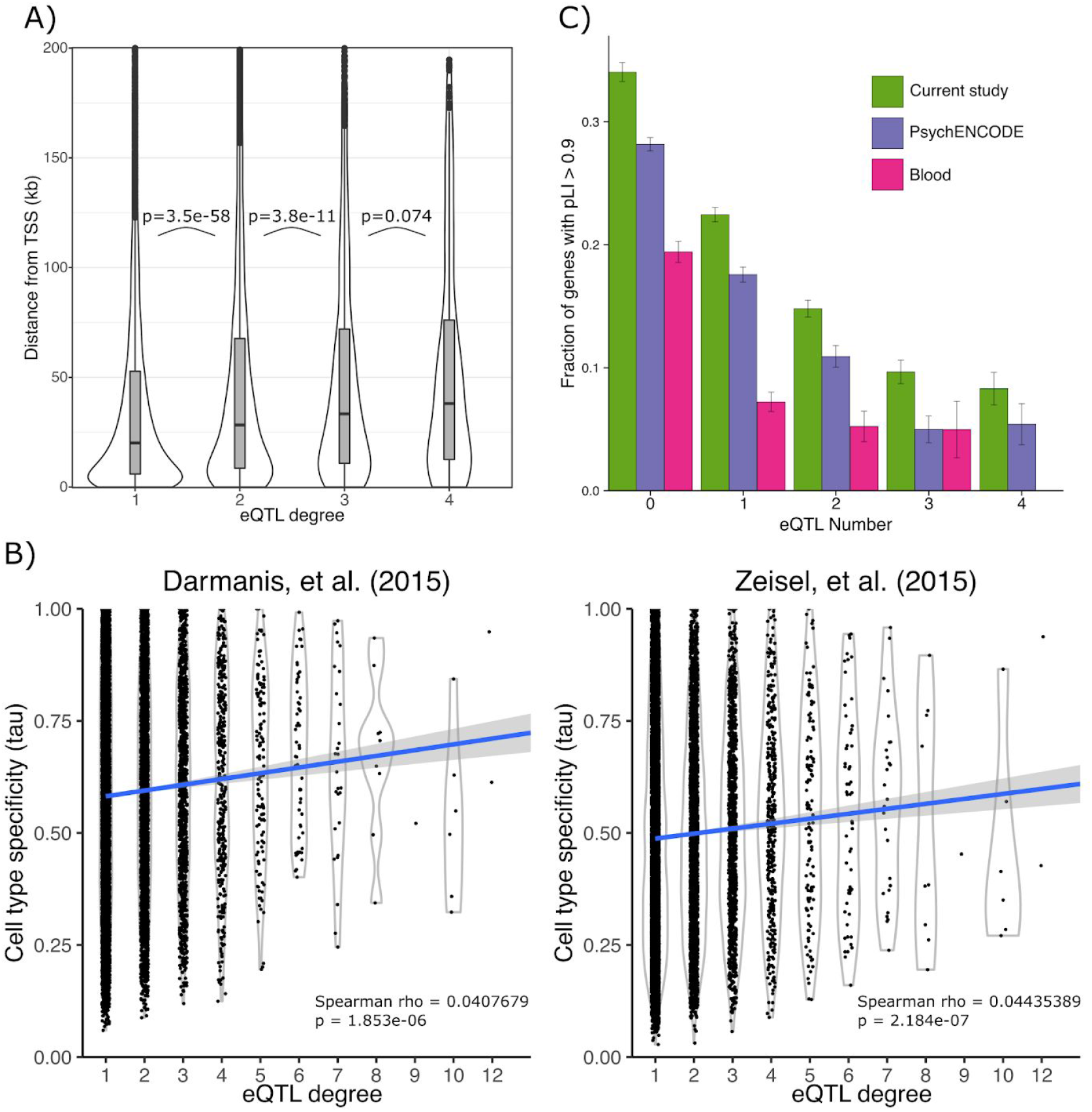
Properties of conditional eQTLs. **A)** The distribution of the distance to the transcription start site is shown for the lead variant for eQTL analysis of increasing degree. P-values indicate significance of one-sided Wilcoxon test between adjacent groups. Box plot indicates median, interquartile range (IQR) and 1.5*IQR. **B)** Cell type specificity metric tau plotted against the number of independent eQTLs discoverged for each gene. **C)** The fraction of genes with high evolutionary constraint (pLI > 0.9) is shown increasing eQTL degree for the current study, PsychENODE (Wang et al., 2018a), and whole blood (Glassberg et al., 2019). Error bars indicate standard error based on asymptotic estimate of binomial proportion.

**Supplementary Figure 3:**
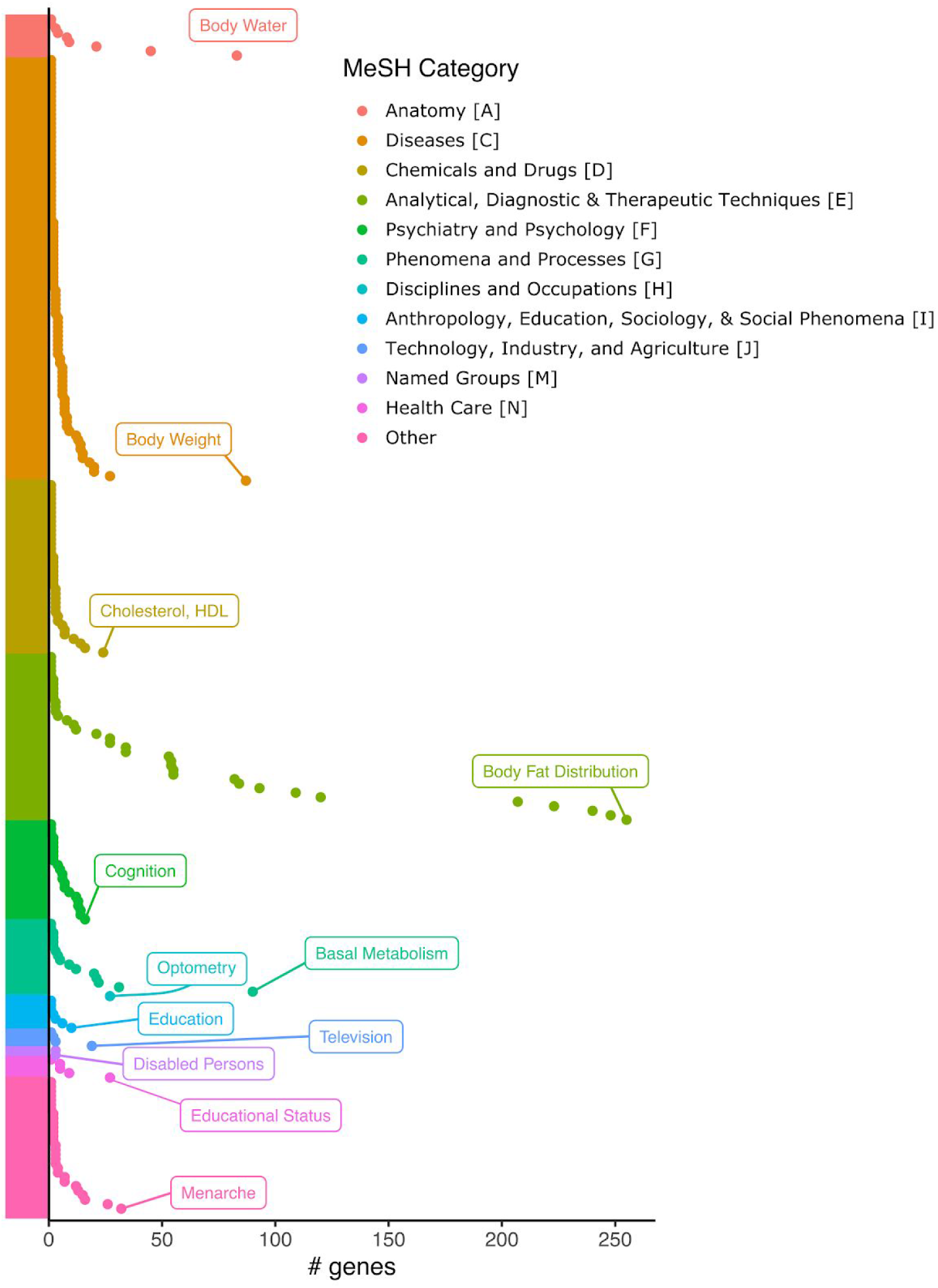
Number of genes colocalizing for each MeSH category with CLPP > 0.01. The phenotype with the highest number of colocalized genes for each MeSH category is indicated.

**Supplementary Figure 4:**
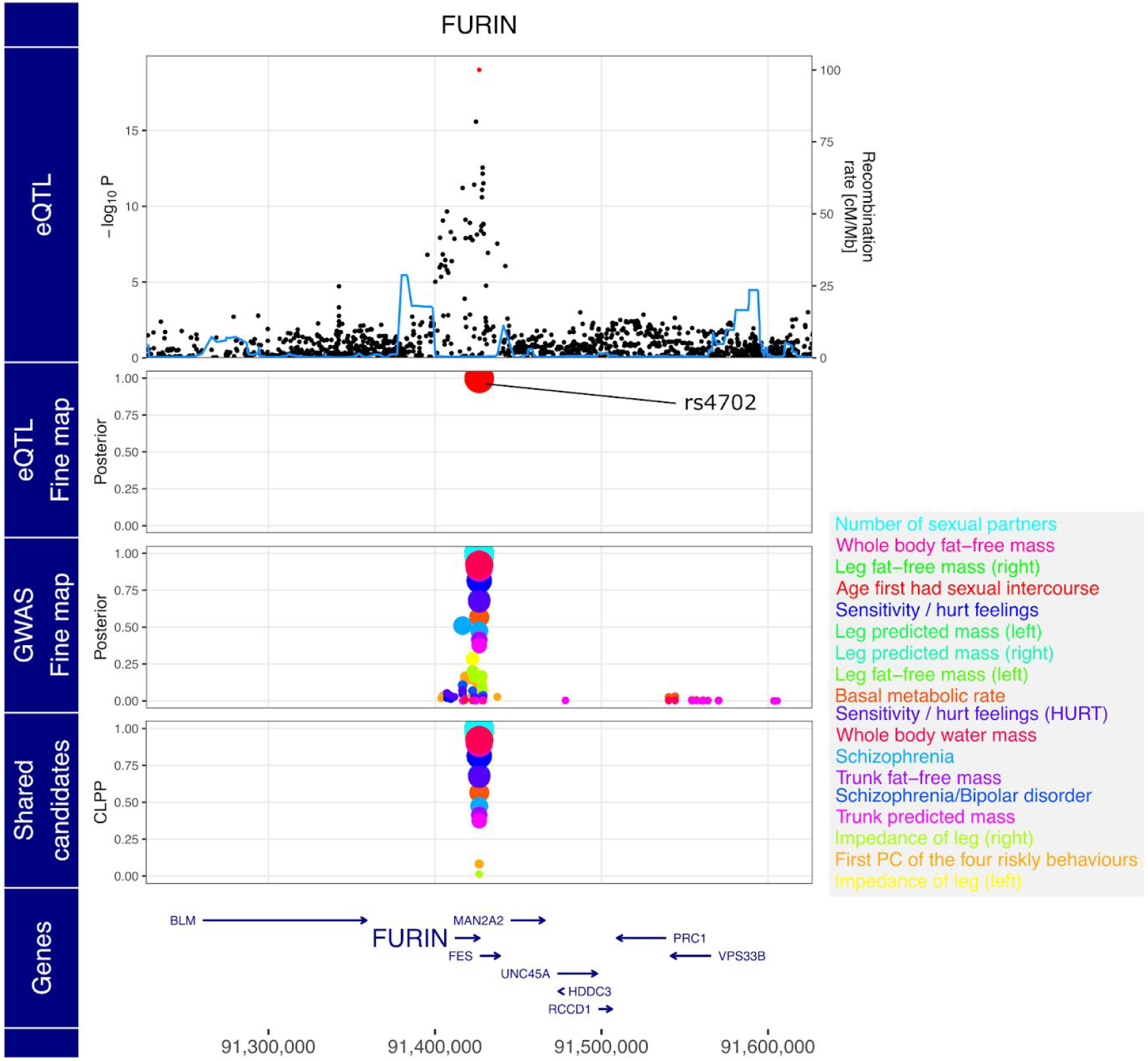
Expression of FURIN and risk for multiple complex traits share rs4702 as a candidate causal variant. Starting from the top, the plot shows -log_10_ p-values from eQTL analysis, poster probabilities from statistical fine-mapping of eQTL results, poster probabilities from statistical fine-mapping of GWAS results, and colocalization posterior probabilities (CLPP) for combining eQTL and GWAS fine-mapping. Traits are shown in the box on the right in decreasing order to CLPP value.

## Supplementary Tables

**Supplementary Table 1.** Trait/Disease, abbreviation and reference for GWAS included in LD-score regression analysis.

| **Abbreviation** | **Trait/disease** | **Reference (doi)** |
| --- | --- | --- |
| ADHD | Attention deficit hyperactivity disorder | 10.1016/j.jaac.2010.06.008 |
| ALS | Amyotrophic lateral sclerosis | 10.1016/j.neuron.2018.02.027 |
| PD | Parkinson's disease | 10.1016/S1474-4422(19)30320-5 |
| RA | Rheumatoid arthritis | 10.1038/nature12873 |
| BMI | Body mass index | 10.1038/nature14177 |
| EduYear | Educational years | 10.1038/nature17671 |
| HEIGHT | Height | 10.1038/ng.3097 |
| CD | Crohn's disease | 10.1038/ng.3359 |
| IBD | Inflammatory bowel disease | 10.1038/ng.3359 |
| UC | Ulcerative colitis | 10.1038/ng.3359 |
| CAD | Cardiovascular disease | 10.1038/ng.3396 |
| DS | Depression symptom | 10.1038/ng.3552 |
| Neu | Neuroticism | 10.1038/ng.3552 |
| SLE | Systemic lupus erythematosus | 10.1038/ng.3603 |
| T2D | Type 2 Diabetes | 10.1038/s41588-018-0241-6 |
| DRINKING | Alcohol Assumption | 10.1038/s41588-018-0309-3 |
| AD | Alzheimer's disease | 10.1038/s41588-018-0311-9 |
| ASD | Autism spectrum disorder | 10.1038/s41588-019-0344-8 |
| BD | Bipolar disorder | 10.1038/s41588-019-0397-8 |
| MDD | Major depression disorder | 10.1038/s41593-018-0326-7 |
| SZ | Schizophrenia | 10.1101/2020.09.12.20192922 |
| MS | Multiple sclerosis | 10.1126/science.aav7188 |

## Notes

### Competing Interest Statement

The authors have declared no competing interest.

### Author Declarations

Data used for this analysis comes from the AD Knowledge Portal (https://adknowledgeportal.synapse.org) and PsychENCODE Knowledge Portal (https://www.synapse.org/#!Synapse:syn4921369) and is hosted on the Sage Bionetworks Synapse platform for access by qualified investigators. Data was generated from post-mortem tissue and has been de-identified according to the Synapse terms of use, and is available through the submission of a AD Knowledge Portal Data Use Certificate (https://adknowledgeportal.synapse.org/DataAccess/DataUseCertificates) or by application to the NIMH Repository and Genomics Resources (https://psychencode.synapse.org/DataAccess), respectively. GTEx data users here is publicly available at https://www.gtexportal.org

